# Flattening the Curve is Imperative: When China Relaxes the Dynamic Zero COVID-19 Policy

**DOI:** 10.1101/2022.12.15.22283335

**Authors:** Shilei Zhao, Tong Sha, Yongbiao Xue, Hua Chen

## Abstract

Since December 7, 2022, China relaxed the strict dynamic zero COVID-19 policy. Here we quantitatively analyze its potential impacts on COVID-19 trend with the epidemiological model SUVQC using the population and parameter settings of Beijing as a case. Our results indicate that if non-pharmacological interventions are completely ceased, the ICU bed demand number will peak in 26 days at ∼23.88 thousand, which is 18 times the total number of ICU beds in Beijing. COVID-19 Omicron will cause 31,817 deaths in Beijing in the first year. We urge that the flattening curve strategy is necessary to slow down the infection and avoid overwhelming the healthcare system.

---

In the past three years, China had been implementing the dynamic zero policy to control the COVID-19 epidemic. On December 7, 2022, China National Health Commission retracted the dynamic zero policy by announcing the Ten New Covid-19 Rules. Most cities in China ceased strict local measures since then. In Beijing, the regular nucleic acid (RT-PCR) testing and the health code checking are not required anymore; and tested positive patients with mild symptoms can be self-quarantined at home. However, the current vaccination rate (proportion of booster doses administered) in China is only 56.9% (https://ourworldindata.org/covid-vaccinations, Our World in Data),^1^ most of which are inactivated vaccines with lower protection efficacy. The pandemic trend is unclear under the drastic swing of containment policy, although Omicron causes much milder symptoms. Here we quantitatively assess the potential impact of the new measures. We analyze the level of healthcare system being overwhelmed after retracting the dynamic zero policy, and compare the number of ICU beds demanded and available in Beijing.

We use the SUVQC model (see details in the supplementary information) to estimate the number of ICU bed needs with the following parameter settings.^2, 3^ The population size of Beijing is *N* = 21,893,000. Since the efficacy of inactivated vaccines to protect infection of Omicron is very limited,^4^ we set *V*(t)=0, *U*_*V*_(t)=0, *η*(t)=0, *λ* _1_=1, *λ* _2_=0. The basic reproductive number (R_0_) of Omicron is set to be 9.5 following previous studies (the R_0_ of Omicron BF.7 lineage prevalent in Beijing may be higher than 9.5);^5^ the generations time is *g*=2.2 days ( *α* = log(*R*_0_ ) / *g* = 1.0233 *γ*_1_ = *α* / *R*_0_ = 0.1077 );^6^ hospitalizations and ICU admissions are 2.6% ^7^ and 0.27% (the number is chosen based on the estimation of risk of developing severe/critical disease in Shanghai,^8^ smaller than the estimation of 0.47% in the US^7^) for Omicron patients respectively. The mean ICU bed occupation time is estimated to be 12 days per individual based on the daily ICU admissions for COVID-19 in the US (https://ourworldindata.org/covid-hospitalizations, Our World in Data), the proportion of patients who require ICU care, and the daily number of new cases in the US ( *γ* _3_ = 1/12 ). Assume that the duration of immunity against COVID-19 is one year after the infection and with 100% protection efficacy ( *t*_*s*_ = 365 ). We set *U*_S_(0) (the number of initial infected individuals who are infectious and unquarantined) to be 200, identical to the number of infected from social screening reported by Beijing Municipal Health Commission in December 7, 2022 (note that the real number of unquarantined infected individuals may be much higher that the reported). The number of *C*(0) (the number of initial cumulative confirmed infected cases) is set to be 19,588 (https://voice.baidu.com/act/newpneumonia/newpneumonia/). We set the number of initial dead individuals *D*_C_(0)=0, the number of initial recovered individuals *R*_C_(0)=0, the number of initial susceptible individuals *S*(0)=*N*-*C*(0), the initial number of quarantined, infected individuals Q(0)=0, the initial number of active cases *A*(0)=8,539 (https://voice.baidu.com/act/newpneumonia/newpneumonia/). We set *k*_1_=0 and *k*_2_=0.0009 according to the fatality rate of Omicron patients in Shanghai^8^. The number of ICU beds occupied is 0.0027*A*(t).

Our analysis indicates that the ICU bed demand will peak at ∼23.88 thousand, around 26 days after relaxing the dynamic zero policy, which is 18 times the total number of available ICU beds in Beijing. Considering we may underestimate the basic reproductive number and the initial unquarantined infected number, the peak time of ICU bed demand may be earlier than the estimation with higher peak value. After several waves of ups and downs, the number of active cases and ICU bed demand will reach an equilibrium state. At this state, the daily new confirmed cases will be around 50,000 and the ICU bed demand is around 1,617; even if all ICU beds are available for the treatment of COVID-19 patients, they still can only cover 81.76% of the demands (By the end of 2021, the total number of ICU beds in Beijing is 1322, and the ICU per 100,000 people is 6.04,^9^ which is only 17.40% of that in the United States, https://ourworldindata.org/, Fig. 1A).

**Fig. 1.**
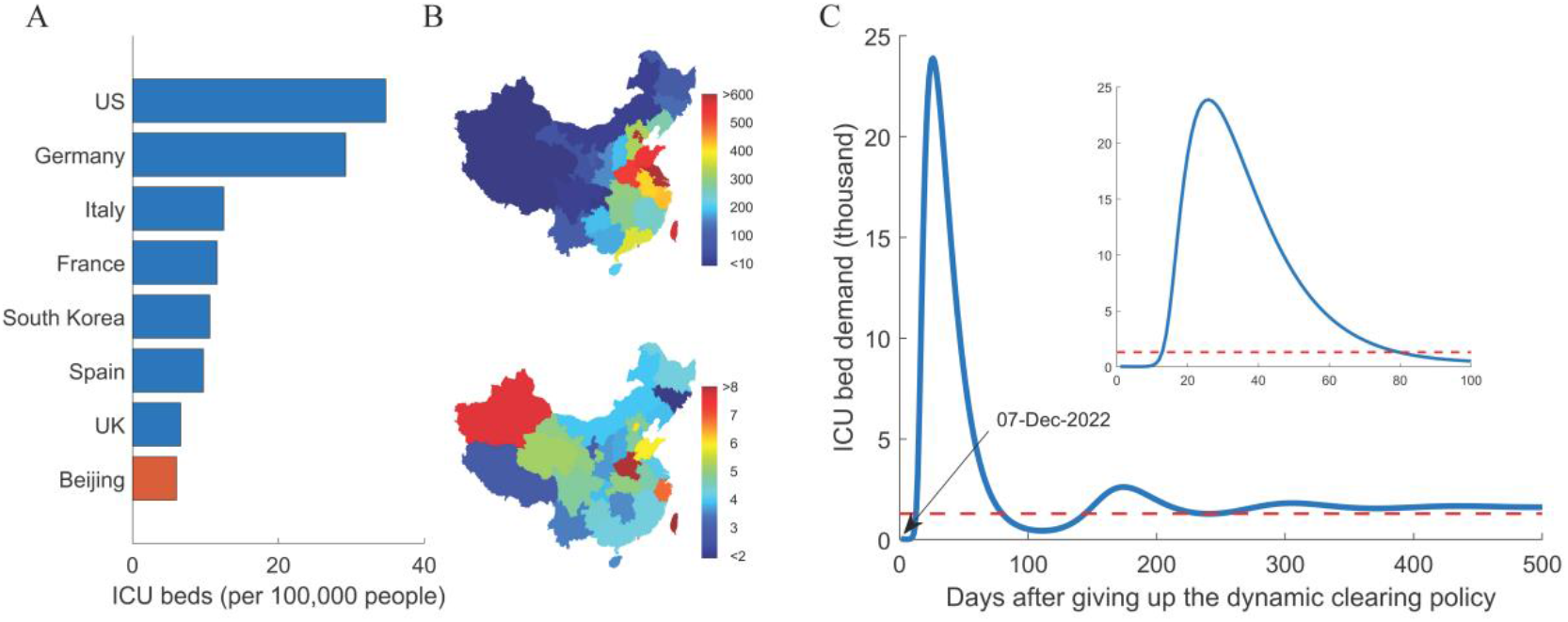
Healthcare system overwhelmed by the shortage of medical resources after retracting the dynamic clearing policy. (**A**) ICU beds per 100,000 people compared with the developed countries. (**B**) Uneven distribution of population and medical resources in China. The colors on the below map represent population density and the colors on the bottom map represent the number of ICU beds per 100,000 people. (**C**) Prediction of the ICU bed demand using SUVQC model after retracting the dynamic zero policy. We used the hospitalizations rate of 2.6% and the ICU admissions rate of 0.27% for Omicron patients. The red line indicates the total number of ICU beds in Beijing.

The peak number of active cases and hospitalizations will reach 8.85 million and 0.23 million respectively. The equilibrium values of these two numbers will be 0.5990 million and 0.0156 million. By assuming a fatality ratio of 0.09% under situations without overwhelming the healthcare system,^7^ it will cause 31,817 deaths in the first year, and ∼16,397 deaths annually in the following years. According to previous studies, the average number of hospitalization days due to influenza was 8.6199 days; the number of hospitalizations due to lower respiratory tract infection of influenza in China was 739,000;^10^ and the total hospitalization days were 6.37 million days. On average, 52,400 people occupy hospital resources every day during the flu season. For Beijing, the influenza burden for hospital is around 819 per day in proportion of population. The hospitalizations caused by Omicron infection will be 19.02 times that caused by influenza during flu season.

To avoid overwhelming the healthcare system at the equilibrium state in Beijing, we need ∼1617 ICU beds for SARS-CoV-2 patients, and more professional doctors and nurses. We must flatten the curve of the first wave of the pandemic after relaxing the dynamic zero policy (Fig. 2). The flattening curve strategy is different from the dynamic zero policy, which aims to restrict the spread of the virus, and the herd immunity strategy of complete liberalization. Flattening curve strategy is to keep the rate of virus spreading low enough with some level of non-pharmacological interventions (NPI). The goal is to diminish the spread of the epidemic to avoid straining the health care system so that severely ill patients can receive medical treatment; and put medical resources at the maximum load of treating new coronary pneumonia patients such that the population can achieve herd immunity at the fastest speed while under the premise of the lowest number of deaths. For flattening curve strategy, R_t_ is still > 1, and the epidemic spreads exponentially but with a lower rate. In addition, the economic loss is smaller in the short term than the rigorous interventions under dynamic zero policy.

**Fig. 2.**
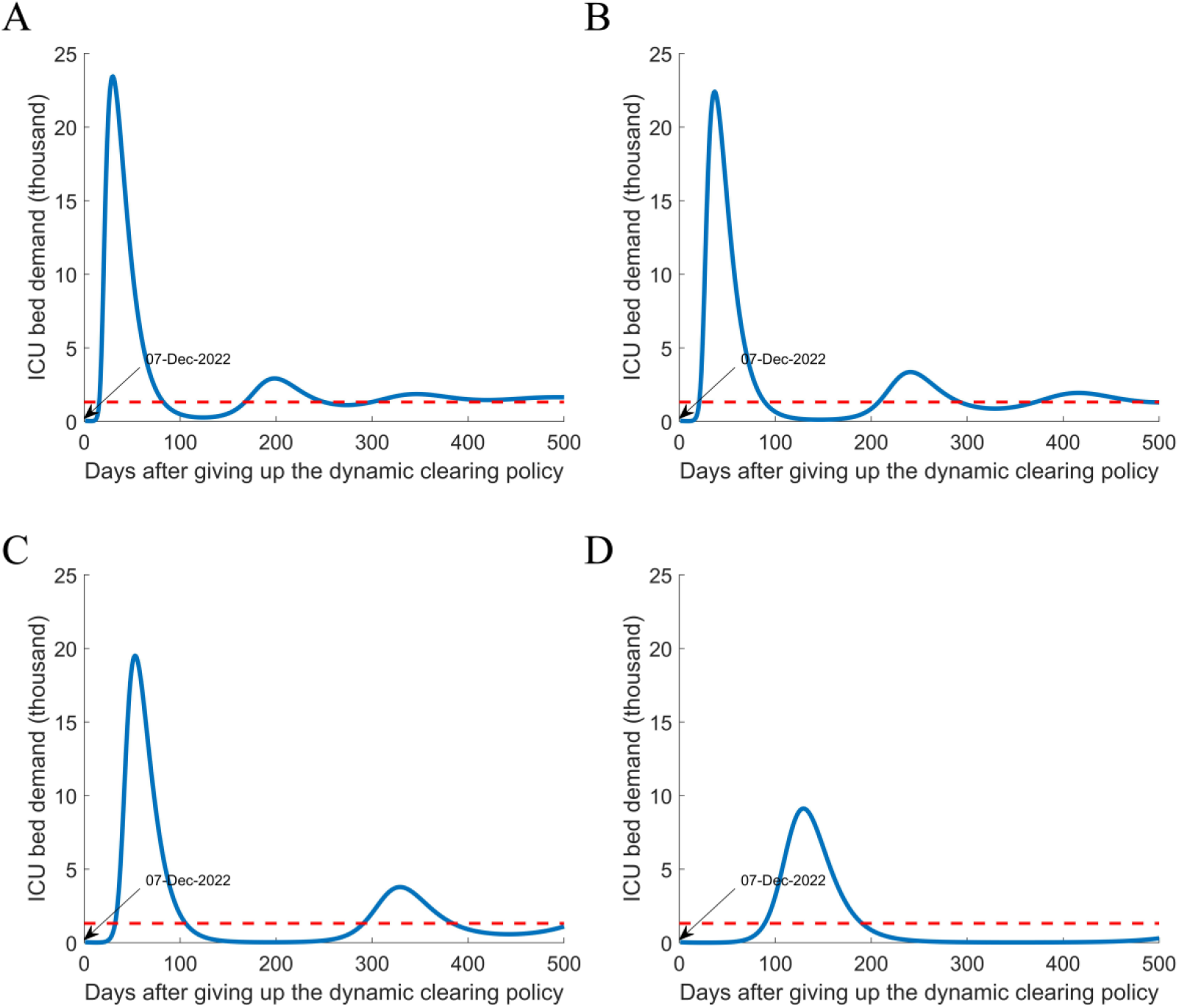
Estimation of ICU demand under different NPI intensities of the flattening curve strategy. (**A-D**): 80%, 60%, 40%, and 20% of transmissions of the usual level. The peak values are 23444, 22414, 19511, and 9124 respectively.

Note that the distribution of medical resources in China is extremely uneven (Fig. 1B) and the bed utilization rate is generally higher than 60%. The situation in areas outside Beijing will be even worse. It is necessary to adopt the flattening curve strategy, and in the meanwhile, accelerate the development of drugs for hospitalized patients and vaccines with more protection efficacy.

To implement the flattening curve strategy, we urge increasing the vaccination rate of the elderly to reduce deaths and relieve medical pressure. Higher levels of NPI, including, social distancing, wearing masks, indoor ventilation and air filtering, regular rapid antigen tests (RAT) and self-examination, and restricted access to public space for those tested positive with RAT, are still very necessary to slow the spread of the virus.

## Data Availability

All data produced are available online at

https://voice.baidu.com/act/newpneumonia/newpneumonia/

https://ourworldindata.org/covid-vaccinations

## Author contributions

SZ and TS did statistical analysis; SZ and HC wrote the manuscript; HC and YX supervised the study.

## Acknowledgments

The work was supported by the National Key R&D Program of China (Grant No.2021YFC0863400).

## Supplementary information

The Susceptible-Unquarantined-Vaccinated-Quarantined-Confirmed (SUVQC) is an epidemiological model to quantitatively analyze and predict the epidemic dynamics of COVID-19. It is an ordinary differential equation model with the following form:

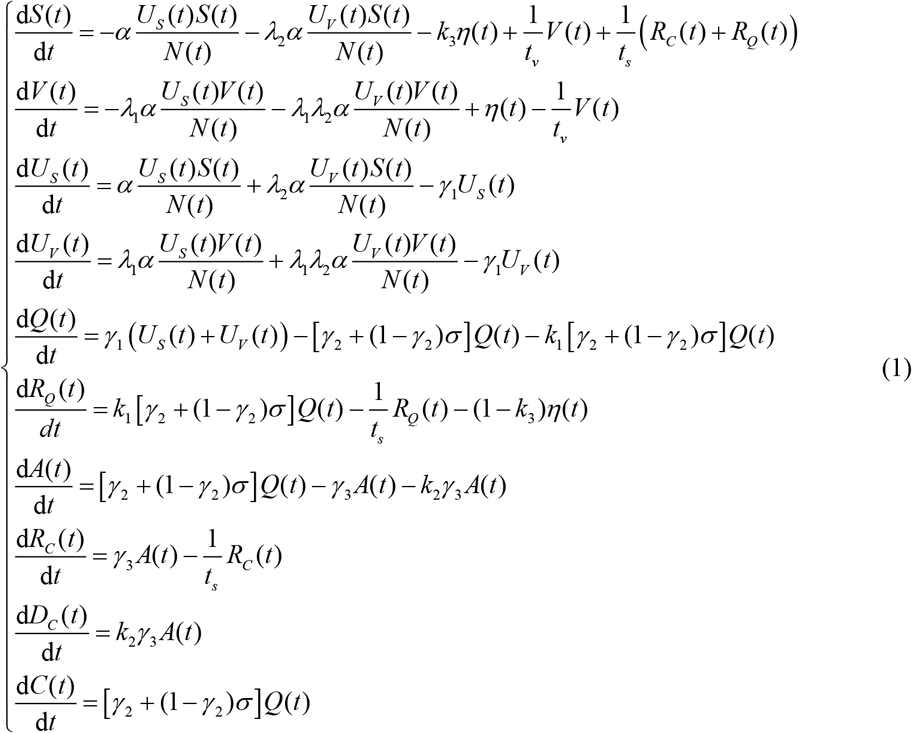

Please see details of variables and parameters in the previous study.^1^

